# Mental Health, Substance Use, and the Importance of Religion during the COVID-19 Pandemic

**DOI:** 10.1101/2022.11.29.22282907

**Authors:** Ji-Yeun Park, Thushara Galbadage, Hyuna Lee, David C. Wang, Brent M. Peterson

**Author notes:** Corresponding Authors: Ji-Yeun Park and Thushara Galbadage.

## Abstract

COVID-19 has impacted all areas of life, with lasting effects on physical, mental, and societal health. Specifically, COVID and related losses have exacerbated prolonged grief responses and mental disorders including depression and anxiety. These mental health concerns are associated with increased detrimental coping strategies including substance use. In this study, we analyzed secondary data from the National Survey on Drug Use and Health (NSDUH) collected during the COVID-19 pandemic. Our results showed a positive association between serious psychological distress and marijuana use, while frequent religious service attendance acted as a moderator in this relationship. Individuals involved in communal religious activity were less likely to use marijuana. This study highlights the impact of religion and faith in bringing hope and purpose during periods of loss, coping with stress, grief, mental health challenges, and substance use.

## Introduction

Nearly three years after the onset of the COVID-19 pandemic, nations and communities have struggled to return to a sense of normality from the devastating impacts of the SARS-CoV-2 virus. At a time when many nations are rescinding large-scale public health orders and restrictions, some even declaring an end to the COVID-19 pandemic altogether, the long-term implications of the pandemic have been largely overlooked. Regardless of the rationale, whether that be overall fatigue, a reticence to address controversial issues, or issues not yet identified, it does appear that a wave of potential public health crises currently exists, and may persist well into the foreseeable future.

### COVID-19 pandemic and grief response

Much like other viral outbreaks and natural disasters, COVID-19 will likely be associated with prolonged grief responses or disorders in affected communities (Eisma, Boelen, & Lenferink, 2020). These responses are either primary or secondary and can be attributed to grief for the loss of former self-identity and lifestyle, bereavement, or grief for the changed world post-pandemic (Verdery, Smith-Greenaway, Margolis, & Daw, 2020). With greater than one million lives lost during the pandemic, many of these deaths occurring in isolation This had a significant impact on the dying and grieving process for both the patient and their loved ones (Galbadage, Peterson, Wang, Wang, & Gunasekera, 2020). Rites of passage and rituals such as these are examples of deep, meaningful, and emotionally charged human experiences. The disruption or loss of integral experiences associated with the death and burial of loved ones has forced people to find ways to cope with stress and unresolved tensions (Corpuz, 2021). Consequently, longer-term bereavement among loved ones who lost their relatives may have negatively impacted an individual’s ability to fully experience and navigate the various stages of grieving: denial, anger, bargaining, depression, and acceptance (Gonçalves Júnior, Moreira, & Rolim Neto, 2020).

### Mental health & societal impact of COVID

COVID-19 pandemic-related mental health and substance use disorders (SUD) have risen in incidence and prevalence, exacerbating an already elevated mental health crisis in the United States (U.S.) and beyond. Individuals who have experienced pandemic-related stressors including acute and chronic illness, the loss of loved ones, and financial strain, may experience increased fear, anxiety, anger, and depression as responses to stressors (Levine, 2022). Systems-based frameworks which have received empirical support have highlighted the interrelatedness of physical health, financial health, mental health, and spiritual well-being (She, Wang, Canada, & Poston, 2022; K. Smith et al., 2022). In the Household Pulse Survey, 42.6% of U.S. adults reported feelings of anxiety and depression in 2020 and these rates have remained high throughout 2021 and 2022 (CDC, 2022b). In contrast, only about 9.5-to-11.7% of U.S. adults reported debilitating feelings of anxiety and depression in 2019 (CDC, 2019). This represents an approximately four-fold increase in identified anxiety and depression disorders as a result of the COVID-19 pandemic, which could persist long into the foreseeable future. Consequently, lingering mental health struggles and pandemic-related stresses have attributed to increased drug-related overdose deaths resulting in 107,000 deaths in 2021-2022 in the U.S. alone. In contrast, before the pandemic, drug overdose deaths in 2019 in the U.S. were around 68,000 (Ahmad, Cisewski, Rossen, & Sutton, 2022).

Human interaction and experiential learning may lead to positive stress-related coping mechanisms, the development of life lessons, and quality of life improvements. The pandemic has, essentially, been a generational mental health crash course by forcing people to learn to cope with multiple stresses occurring simultaneously without many of the social and emotional resources typically available to them. Positive coping skills can help individuals, their loved ones, and their community increase resilience. Furthermore, when positive adaptive coping tools are unavailable, vulnerability to maladaptive behaviors including increased consumption of alcohol, illegal or recreational drug use like marijuana, heroin, cocaine, methamphetamines, and misuse of prescription drugs like opioids increases (CDC, 2022a).

### Substance use & mental health disorders

There is a bidirectional relationship between substance use and psychological distress (Womack, Shaw, Weaver, & Forbes, 2016). For example, marijuana use and distress can reciprocally exacerbate each other over time. One of the common reasons people use marijuana is to deal with their stress and anxiety as a coping strategy. The inadequate ability to tolerate psychological distress may result in maladaptive coping methods through the use of marijuana during times of heightened negative emotions. Conversely, frequent marijuana use is often associated with increased psychological distress and depressive symptoms as a result of the dulling effects of the drug on feelings and emotions (Khadrawy, Sawie, Abdel-Salam, & Hosny, 2017; Rhew, Cadigan, & Lee, 2021). As a mental health disorder, SUD affects an individual’s behavior resulting in an inability to control the use of legal or illegal drugs (Hasin et al., 2013). Patients diagnosed with SUD are likely to also be diagnosed with additional mental health disorders, with co-morbid SUD and mental health disorders more common than SUD alone (Ross & Peselow, 2012). For example, studies have indicated that high rates of SUD have been observed concomitantly with anxiety disorders or depressive states (Conway, Compton, Stinson, & Grant, 2006; Wolitzky-Taylor, Operskalski, Ries, Craske, & Roy-Byrne, 2011). During the COVID-19 pandemic, an increase in comorbid SUD and mental health disorders were observed (Johnson, 2021). While substance abuse is often used in response to stress and grief, it is maladaptive because it ultimately complicates and hinders adaptive grief and stress responses. This dynamic has been at play throughout the COVID-19 pandemic (Caparrós & Masferrer, 2020).

### Religion, Coping, and Distress during the COVID-19 pandemic

Recent research has highlighted the multifold means by which religion has been implicated in the meaning-making and shaping of one’s experiences during the COVID-19 pandemic, with notable consequences on mental health and well-being. To illustrate, a recent study explored the implications of divine and demonic attributions towards COVID-19 vaccines and found that demonic (but not divine) attributions were linked with not only anti-vaccination attitudes and lower odds of vaccination, but also greater interpersonal anger, anxiety, depression, and spiritual struggles (Exline, Pait, Wilt, & Schutt, 2022). Speaking of the latter, religious and spiritual struggles (e.g., feeling punished or abandoned by God, uncertain of one’s faith beliefs, feelings of strain with members of one’s spiritual community) were found to be a common experience among many Americans during the COVID-19 pandemic and such struggles were in turn associated with negative mental health outcomes along with lower self-rated physical health outcomes (Upenieks, 2022). Religious practices such as prayer also bear implications on mental health longitudinally during the COVID-19 pandemic (Lowe, Wang, & Chin, 2022); certain prayer practices such as meditative and colloquial prayer (but not petitionary prayer) predicted lower anxiety, depression, and burnout because they provided a means by which an individual can proactively and adaptively approach (instead of avoid) their experiences and circumstances during the pandemic.

While it is believed that COVID can lead to an exacerbation of mental health disorders and an associated increase in substance use, there is a knowledge gap in moderating factors that can moderate the negative relationship between mental illness and substance use during the COVID-19 pandemic. Therefore, the rationale for this comparative data assessment was to investigate potential associations between pandemic-associated mental health diseases, the use of substances as a mechanism to alleviate negative aspects of the experience, and the protective role of religious activities, which for many represents a crucial source of social support and adaptive coping and meaning-making. Specifically, we investigated the degree to which religiosity functioned as a protective factor for substance use among individuals with and without serious psychological distress (SPD) during the COVID-19 pandemic.

## Methods

### Secondary data source

In this study, we analyzed data obtained from the 2020 National Survey on Drug Use and Health (NSDUH). Data are composed of comprehensive face-to-face household interviews of non-institutionalized individuals aged 12 or older. Excluded interviewees included individuals incarcerated in prison, members of the active-duty military, residents of long-term medical care, and people without housing and not living in shelters. NSDUH is a primary source of information on the use of tobacco, alcohol, illegal drug use, and mental illness in the U.S.

Starting in 2020, NSDUH began to use both in-person interviews and web-based interviews. NSDUH data were only collected in Quarters 1 (January to March) and 4 (October to December) of 2020. Most Quarter 1 data were collected via in-person interviews, whereas online data were dominant in Quarter 4 with limited in-person data collection (SAMHSA, 2021). In 2020, NSDUH collected data from 36,284 respondents aged 12 or older. NSDUH is publicly available, de-identified data, and hence did not require IRB approval.

### Study variables

#### Current marijuana use (CMU)

The main dependent variable utilized in this investigation was CMU, highlighted by NSDUH data. Respondents who indicated marijuana use in the past 30 days were considered current users. Since marijuana is the most commonly used illegal drug, it was used as a representation of overall substance use following the Substance Abuse and Mental Health Services Administration (SAMHSA) guidelines.

#### Past-month serious psychological distress

The NSDUH incorporates the Kessler-6 (K6) screening instrument (questionnaire) to assess serious psychological distress (SPD) among respondents aged 18 or older only via a 5-point Likert scale. Scores greater than 13 were categorized as having past-month SPD, whereas scores less than 13 were categorized as not having past-month SPD.

#### Religiosity

The NSDUH measured the three components of religiosity: frequency of religious service attendance, the importance of religious belief, and the influence of religious belief on decision-making. Religious service attendance represents the external aspect of religiosity, whereas religious beliefs represent the internal aspect. In general, people are highly religious when they frequently attend religious services and follow religious beliefs. Two religious variables, the frequency of religious service attendance and the influence of religious belief on decision-making, were included in the analysis because they were found to be negatively associated with substance use (Becerra, Becerra, Gerdine, & Banta, 2014; Edlund et al., 2010). For example, an individual’s decision not to use a substance may be influenced by religious belief that substance use is explicitly prohibited by religious doctrine (Stewart, 2001). Furthermore, a lack of congruence between the internal and external aspects of religiosity is associated with increased odds of substance use (Marsiglia, Ayers, & Hoffman, 2012).

Frequency of religious service attendance was measured by asking “During the past 12 months, how many times did you attend religious services? Please do not include special occasions such as weddings, funerals, or other special events in your answer.” Possible response options were 0 times, 1 to 2 times, 3 to 5 times, 6 to 24 times, 25 to 52 times, and more than 52 times.

Responses were categorized into three levels: (1) 0 times, (2) 1 to 24 times, and (3) more than 25 times. The influence of religious belief on decision-making was assessed with the following question: “My religious beliefs influence my decisions.” Possible response options were strongly disagreed, disagree, agree, and strongly disagree. Responses were categorized into two levels: (1) strongly disagree/disagree and (2) agree/strongly agree.

#### Sociodemographic characteristics

Selected sociodemographic characteristics were included in the analysis as covariates, including sex (male and female), age (18 - 25, 26 - 34, 35 - 49, and 50 +), education (less than high school, high school graduate, some college, and college graduate), and employment status in the past week (full-time, part-time, unemployed/on layoff or looking for work, and others). Race/ethnicity was recorded into 4 levels (non-Hispanic NH-White, NH-Black, Hispanic, and other). Due to the small sample size, we combined NH-Asian, NH-Native American/Alaska Natives, NH-Native Hawaiian/Other Pacific Islanders, and people with multiple races into one group.

#### Statistical Analysis

Descriptive statistics were conducted to examine sample characteristics. We then estimated the prevalence of CMU by SPD status and then the prevalence of CMU by religious factors. Next, we performed the multivariate logistic regression, adjusting for all other covariates including sex, age, race/ethnicity, education, and employment status. The multivariate logistic regression was conducted to examine factors associated with marijuana use during the COVID-19 pandemic. Interaction terms were then added to the model to test for any moderating effect of religiosity. The NSDUH sampling weights were adjusted for all analyses, to account for the complexities of the survey. All analyses were conducted using STATA Version 17 (Stata, College Station, TX).

## Results

### Sample characteristics

Table 1 provides the overall characteristics of the adult sample from the 2020 NSDUH (*N* = 27,170). The majority of the study sample was composed of adults aged 50 years of age or older (46.1%), and NH-White (62.7%), and maintained a full-time employment status (43.1%). In 2020, according to the NSDUH survey results, approximately 7.0% of U.S. adults had SPD based on the Kessler-6 score in the past month. Approximately, 49.1% of the survey respondents reported that they did not attend any religious services or participate in any religious activities. Respondents (29.5%) also indicated that they were infrequent attendants to religious services (1 - 24 times per year) and 21.4% reported they were frequent attendants (25 times per year or more). Yet, in this sample, 67.0% of respondents reported that their religious belief influenced their behavioral decisions.

**Table 1.** Sample Characteristics from NSDUH 2020 (N = 27,170)

| **Variable** | **N** | **% [95% CI]** |
| --- | --- | --- |
| **Sex:** |  |  |
| Male | 12185 | 48.3 [47.2-49.4] |
| Female | 14985 | 51.7 [50.6-52.8] |
| **Age:** |  |  |
| 18-25 | 7946 | 13.3 [12.7-13.9] |
| 26-34 | 5711 | 16.1 [15.4-16.8] |
| 35-49 | 7418 | 24.6 [23.7-25.5] |
| 50+ | 6095 | 46.1 [44.7-47.4] |
| **Race/ethnicity:** |  |  |
| NH-White | 18144 | 62.7 [61.1-64.2] |
| NH-Black | 2437 | 12.1 [11.1-13.2] |
| Hispanic | 3728 | 16.7 [15.5-18.0] |
| Others | 2861 | 8.51 [7.94-9.12] |
| **Education:** |  |  |
| Less than high school | 2244 | 11.3 [10.5-12.3] |
| High school graduate | 5683 | 27.7 [26.5-29.0] |
| Some college | 8371 | 30.5 [29.5-31.5] |
| College graduate | 10872 | 30.5 [29.2-31.7] |
| **Employment status in the past week:** |  |  |
| Worked at a full-time job | 12569 | 43.1 [42.1-44.2] |
| Worked at a part-time job | 3333 | 10.5 [9.9-11.0] |
| Unemployed/on layoff, looking for work | 1140 | 3.9 [3.4-4.3] |
| Others | 8835 | 42.6 [41.3-43.8] |
| **Past-month SPD** |  |  |
| No | 24443 | 92.8 [92.3-93.3] |
| Yes | 2727 | 7.2 [6.7-7.7] |
| **Frequency of religious service attendance in the past year** |  |  |
| 0 time | 13604 | 49.1 [47.9-50.3] |
| 1-24 times | 7689 | 29.5 [28.5-30.4] |
| More than 25 times | 5023 | 21.4 [20.3-22.6] |
| **My religious beliefs influence my decisions** |  |  |
| Disagree | 10191 | 33.0 [31.8-34.2] |
| Agree | 15884 | 67.0 [65.8-68.2] |

### The presence of SPD & association with CMU

The prevalence of CMU (within the past 30 days) among U.S. adults was 12.1% in 2020. CMU had a higher prevalence among people with SPD (27.2%) compared to those without (10.9%) (Figure 1 a). Similar patterns were observed in the lifetime and past-year prevalence of marijuana use. In 2020, 18.2% of U.S. adults indicated marijuana use in the past year. By SPD status, those with SPD (39.8%) were more likely to report past-year marijuana use than those without (16.5%) (Figure 1 b). In 2020, among U.S. adults, the lifetime prevalence of marijuana use was 48.4%. Among those with SPD, 68.8% (95% CI = 64.7 - 72.6) reported having ever used marijuana, which was significantly higher than in those without SPD (46.8%, 95% CI = 45.6 - 48.0) (Figure 1 c). Multivariate logistic regression analysis also showed that SPD was a significant factor associated with CMU. Those without SPD had decreased likelihood of being current marijuana users than those with SPD (AOR = 0.48, 95% CI = 0.29 - 0.57) (Table 3).

**Figure 1.**
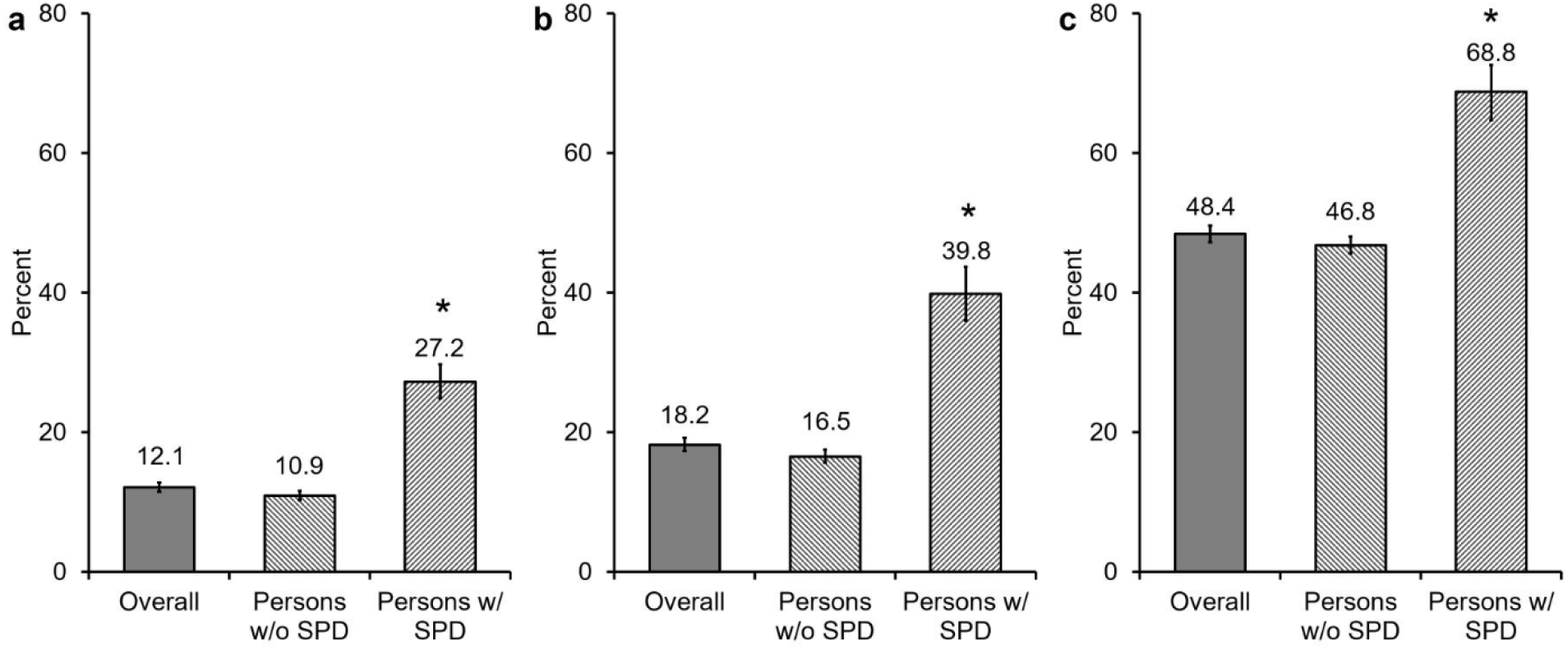
Prevalence of marijuana use comparing persons without (w/o) serious psychological distress (SPD) and persons with (w/) SPD. Secondary data was obtained from the 2020 National Survey on Drug Use and Health (NSDUH) conducted by the Center for Behavioral Health Statistics and Quality within the Substance Abuse and Mental Health Services. NSDUH used the Kessler-6 (K6) screening instrument to assess serious psychological distress (SPD) among respondents aged 18 or older. (a) past-month marijuana use, (b) past-year marijuana use, and (c) ever use of marijuana. Past-month marijuana use indicates CMU status during the COVID-19 pandemic. Error bars represent the upper and lower limits of the 95% confidence intervals (C.I.s) * Statistically significant differences between persons with SPD and persons without SPD: if the confidence intervals are not overlapping, the difference between the value is significantly different.

### Religious service attendance & CMU

The prevalence of CMU decreased with the number of religious service attendance (Table 2). Among people with SPD, the prevalence of CMU was 29.3%, 24.0%, and 22.3% for non-attendants, infrequent attendants, and frequent attendants, respectively. Among people without SPD, the prevalence of CMU was 15.2%, 9.4%, and 2.9% for non-attendants, infrequent attendants, and frequent attendants, respectively.

**Table 2.** Prevalence of CMU by Religious Factors.

| **Variable** | **People with SPD<sup>a</sup>**<br>**(N=2,699)** |  | **People without SPD<sup>a</sup>**<br>**(N=23,376)** |  |
| --- | --- | --- | --- | --- |
|  | **N** | **% [95% CI]** | **N** | **% [95% CI]** |
| **Frequency of religious service attendance in the past year** |  |  |  |  |
| 0 time | 1693 | 29.3 [25.8-33.1] | 11911 | 15.2 [14.1-16.4] |
| 1-24 times | 773 | 24.0 [19.9-28.5] | 6916 | 9.4 [8.3-10.6] |
| More than 25 times | 249 | 22.3 [11.4-39.1] | 4774 | 2.9 [2.2-3.8] |
| **My religious beliefs influence my decisions** |  |  |  |  |
| Disagree | 1407 | 30.9 [27.4-34.6] | 8784 | 17.0 [15.7-18.4] |
| Agree | 1292 | 24.0 [20.4-28.0] | 14592 | 7.7 [6.8-8.7] |

<sup>a</sup> SPD: serious psychological distress

**Table 3.** Results from Multivariate Logistic Regression Predicting CMU Among U.S. Adults.

|  | Main effect model<br>(N=25,382) | Interaction effect<br>model<br>(N=25,382) |
| --- | --- | --- |
| Variable | AOR [95% CI] | AOR [95% CI] |
| **Sex (ref.=male)** |  |  |
| Female | 0.77 [0.67-0.89]** | 0.77 [0.67-0.89]** |
| **Age (ref.=18-25)** |  |  |
| 26-34 | 0.78 [0.65-0.93]** | 0.78 [0.65-0.93]** |
| 35-49 | 0.57 [0.47-0.69]** | 0.57 [0.47-0.69]** |
| 50+ | 0.29 [0.22-0.38]** | 0.29 [0.22-0.38]** |
| **Race/ethnicity (ref.=NH-White)** |  |  |
| NH-Black | 1.21 [0.97-1.49] | 1.21 [0.97-1.49] |
| Hispanic | 0.52 [0.42-0.66]** | 0.52 [0.42-0.66]** |
| Others | 0.61 [0.47-0.78]** | 0.61 [0.47-0.78]** |
| **Education (ref.=Less than high school)** |  |  |
| High school graduate | 1.31 [1.05-1.64]* | 1.31 [1.05-1.64]* |
| Some college | 1.47 [1.17-1.84]** | 1.47 [1.17-1.84]** |
| College graduate | 1.06 [0.81-1.39] | 1.06 [0.81-1.39] |
| **Employment status in the past week (ref.=full time)** |  |  |
| Part-time job | 1.16 [0.97-1.38] | 1.16 [0.97-1.38] |
| Unemployed/on layoff, looking for work | 1.53 [1.11-2.11]** | 1.53 [1.11-2.11]** |
| Others | 1.02 [0.86-1.21] | 1.02 [0.86-1.21] |
| **SPD<sup>a</sup> status (ref.= With SPD<sup>a</sup>)** |  |  |
| Without SPD<sup>a</sup> | 0.48 [0.39-0.57]** | 0.60 [.48-0.77]** |
| **Frequency of religious service attendance in the past year (ref.=0 time)** |  |  |
| 1-24 times | 0.69 [0.58-0.82]** | 0.78 [0.55-1.11] |
| More than 25 times | 0.27 [0.21-0.36]** | 0.87 [0.31-2.44] |
| **My religious beliefs influence my decisions (ref.=disagree)** |  |  |
| Agree | 0.68 [0.56-0.82]** | 0.82 [0.58-1.16] |
| **Frequency of religious service attendance in the past year x SPD<sup>a</sup> status (ref= 0 time, with SPD)** |  |  |
| 1-24 times x without SPD<sup>a</sup> | --- | 0.86 [0.56-1.31] |
| More than 25 times x without SPD<sup>a</sup> | --- | 0.26 [0.08-0.84]* |
| **My religious beliefs influence my decisions x SPD<sup>a</sup> status (ref= disagree, with SPD)** |  |  |
| Agree x without SPD<sup>a</sup> | --- | 0.80 [0.52-1.23] |

<sup>a</sup> SPD: serious psychological distress
Note. AOR=Adjusted Odds Ratio; CI=Confidence Interval.
\*\* < .01, \* < .05

Multivariate logistic regression analysis also showed that religious service attendance played a significant role in marijuana use (Table 3). Both frequent (AOR = 0.27, 95% CI = 0.21 - 0.36) and infrequent (AOR = 0.69, 95% CI = 0.58 - 0.82) religious service attendance was associated with lower odds of CMU compared to those who did not attend religious service (Table 3). Overall, results indicated that individuals involved in some forms of communal religious activity were less likely to use marijuana. Additionally, there was a significant interaction effect between frequent religious service attendance and SPD (AOR = 0.26, 95% CI = 0.08 - 0.84). This suggests that frequent service attendance moderates the relationship between SPD and CMU.

### Influence of religious belief on decision-making & CMU

The prevalence of CMU also varied depending on individuals’ religious beliefs. Among people with SPD, those who reported that religious beliefs would not influence their decisions were more likely to report CMU (30.9%) than those who reported that religious beliefs would influence their decisions (24.0%). Similarly, among people without SPD, CMU was more common among those who reported that religious beliefs would influence their decisions (17.0%) than those who reported that religious beliefs would not influence their decisions (7.7%). In the multivariate logistic regression model, those who reported that religious beliefs would influence their decisions had lower odds of CMU (AOR = 0.68, 95% CI = 0.56 - 0.82) compared with those who reported that religious belief would not influence their decisions. There was no significant interaction effect between the influence of religious belief on decision-making and SPD.

### Age & CMU

Age was negatively associated with marijuana use (Table 3). Those aged 26 - 34 (AOR = 0.78, 95% CI = 0.65 - 0.93), those aged 35 - 49 (AOR = 0.57, 95% CI = 0.47 - 0.69), and those aged 50 + (AOR = 0.29, 95% CI = 0.22 - 0.38) had a lower likelihood of CMU than those aged 18 - 25.

### Additional factors & CMU

Sex, race/ethnicity, education, and employment status were associated with CMU (Table 3). Women were less likely to report CMU than men (AOR = 0.77, 95% CI = 0.67 - 0.89). Compared to NH-White, Hispanics (AOR = 0.52, 95% CI = 0.42 - 0.66) and other racial/ethnic groups (AOR = 0.61, 95% CI = 0.47 - 0.78) were less likely to report CMU. Individuals who completed high school (AOR = 1.31, 95% CI = 1.05 - 1.64) and those who completed some college (AOR = 1.47, 95% CI = 1.17 - 1.84) had increased odds of CMU than individuals with less than a high school diploma. Unemployed individuals also had increased odds of CMU (AOR = 1.53, 95% CI = 1.11 - 2.11) than full-time employees.

### Conclusion

Religion service attendance and the influence of religious belief on decision-making are associated with decreased odds of marijuana use. Given the strong relationship between external and internal forms of religiosity and less substance use, religious communities and faith-based activities may play a significant role in reducing substance use behaviors during and after the COVID-19 pandemic.

## Discussion

COVID-19 impacted our world in many ways. The extensive nature to which COVID-related mental health and accompanying substance use problems are manifesting in the U.S. is just starting to emerge. Understandably, substance use and psychological distress increased during the pandemic. Herein, our findings empirically confirmed this association between substance use and psychological distress during the COVID-19 pandemic. Thus, public health practitioners need to recognize the exigent, yet potentially ominous nature by which these issues could have future implications and develop strategies toward a greater understanding of the influence of long-COVID on aspects of mental health and substance abuse.

### Association between SPD & SUD

Consistent with previous research, our group found a higher prevalence of CMU in people with SPD (Weinberger et al., 2019). Adults with SPD were more likely to report prior month, previous year, and lifetime marijuana use than adults without SPD. Previous studies provided a plausible mechanism linking psychological distress and marijuana use as an emotional management strategy (Aselton, 2012). Another recent study found an association between phycological distress and marijuana use among pregnant women during the COVID-19 outbreak (C. L. Smith et al., 2022). Given the substantial impact of COVID-19 on mental health, our findings are noteworthy. Since the pandemic and its long-term effects are still ongoing, it is important to ensure that mental health screening, referral, and treatment services are made available, and delivered effectively. Especially since mental health interventions aiming to promote positive coping mechanisms and stress management strategies may play a vital role in preventing and reducing marijuana use.

### Religion, faith, mental health, & substance use behavior

The influence of religious belief on decision-making, which is an internal form of religiosity, was an important factor associated with lower marijuana use. This is probably because people who feel that their religious beliefs strongly affect their decision-making tend to be highly integrated within their religious group. Religious affiliation and one’s dedication to faith are closely tied to a person’s source of hope, sense of purpose in life, coping skills, and social support network (Ano & Vasconcelles, 2005; Berthold & Ruch, 2014). Religious beliefs provide a sense of purpose for those that believe in a God and help provide an understanding of role and purpose (Koenig, 2009). The understanding of a greater good and the assurance faith and religion provide for people influence the choices they make including avoidance of substance use behaviors.

This study also found that frequency of religious attendance, which is an external form of religiosity, plays a salient role in preventing marijuana use, which is consistent with findings from a previous study showing that attending religious services was a strong substance use protective factor (Livne et al., 2021). Furthermore, frequent religious attendance was a potential moderator in the relationship between SPD and marijuana use. Practicing faith increases life satisfaction compared to those who have no religious affiliation or do not practice, and can protect against negative emotions that can lead to depression and anxiety (Berthold & Ruch, 2014).

Religious beliefs, practices, and spirituality have been observed to promote mental health through positive religious coping, positive beliefs, and community support as a means to cope with stress and illness (Weber & Pargament, 2014). Coping through stressful life events, such as the pandemic, by religious means can allow one to experience positive psychological outcomes evident through lowered depression and anxiety; while non-religious coping mechanisms can at times lead to greater distress (Ano & Vasconcelles, 2005). These outcomes have bolstered religious service attendance as a means of attenuating pandemic-related stress, and at the same time have disincentivized sustained marijuana use as a possible stress management activity.

### Study limitations

We used cross-sectional NSDUH data in the general population of U.S. adults aged 18 and older. Therefore, our study findings can’t be generalized across other countries. Additional studies conducted in different countries and with longitudinal data are needed to provide more robust evidence of the relationship between SPD and substance use, and the role of faith in this relationship. Furthermore, due to the cross-sectional nature of the data, we could not establish causality between SPD and substance use. Some caution is needed when interpreting our study findings as they indicate only associations. Finally, the NSDUH did not provide information about the religious affiliations of study participants. Hence, we could not specify the relationship between faith, SPD, and substance use by type of religion. Moreover, we are unable to identify the specific mechanisms of change active in religious activity, whether it relates to general social support, meaning-making, or even religious stigma to substance use, for example. Further study is warranted to determine the specific and multi-faceted role that religion might play in the relationship between mental illness and substance use.

## Data Availability

All data produced in the present study are available upon reasonable request to the authors.

## References

Ahmad, F. B., Cisewski, J. A., Rossen, L. M., & Sutton, P. (2022). Provisional Drug Overdose Death Counts. National Center for Health Statistics. Retrieved from https://www.cdc.gov/nchs/nvss/vsrr/drug-overdose-data.htm

Ano, G. G., & Vasconcelles, E. B. (2005). Religious coping and psychological adjustment to stress: a meta-analysis. J Clin Psychol, 61(4), 461–480. doi:10.1002/jclp.20049

Aselton, P. (2012). Sources of stress and coping in American college students who have been diagnosed with depression. J Child Adolesc Psychiatr Nurs, 25(3), 119–123. doi:10.1111/j.1744-6171.2012.00341.x

Becerra, B. J., Becerra, M. B., Gerdine, M. C., & Banta, J. E. (2014). Religion, acculturation, and incarceration: determinants of substance use among Hispanic adults in the United States. J Environ Public Health, 2014, 459596. doi:10.1155/2014/459596

Berthold, A., & Ruch, W. (2014). Satisfaction with life and character strengths of non-religious and religious people: it’s practicing one’s religion that makes the difference. Front Psychol, 5, 876. doi:10.3389/fpsyg.2014.00876

Caparrós, B., & Masferrer, L. (2020). Coping Strategies and Complicated Grief in a Substance Use Disorder Sample. Front Psychol, 11, 624065. doi:10.3389/fpsyg.2020.624065

CDC. (2019). National Health Interview Survey, Estimates of Mental Health Symptomatology by Month of Interview: United States, 2019. National Center for Health Statistics.

CDC. (2022a). Coping With Stress. Retrieved from https://www.cdc.gov/mentalhealth/stress-coping/cope-with-stress/index.html

CDC. (2022b). Long COVID: Household Pulse Survey. June 22, 2022. Retrieved from https://www.cdc.gov/nchs/covid19/pulse/long-covid.htm

Conway, K. P., Compton, W., Stinson, F. S., & Grant, B. F. (2006). Lifetime comorbidity of DSM-IV mood and anxiety disorders and specific drug use disorders: results from the National Epidemiologic Survey on Alcohol and Related Conditions. J Clin Psychiatry, 67(2), 247–257. doi:10.4088/jcp.v67n0211

Corpuz, J. C. G. (2021). Beyond death and afterlife: the complicated process of grief in the time of COVID-19. J Public Health (Oxf), 43(2), e281–e282. doi:10.1093/pubmed/fdaa247

Edlund, M. J., Harris, K. M., Koenig, H. G., Han, X., Sullivan, G., Mattox, R., & Tang, L. (2010). Religiosity and decreased risk of substance use disorders: is the effect mediated by social support or mental health status? Soc Psychiatry Psychiatr Epidemiol, 45(8), 827–836. doi:10.1007/s00127-009-0124-3

Eisma, M. C., Boelen, P. A., & Lenferink, L. I. M. (2020). Prolonged grief disorder following the Coronavirus (COVID-19) pandemic. Psychiatry Res, 288, 113031. doi:10.1016/j.psychres.2020.113031

Exline, J. J., Pait, K. C., Wilt, J. A., & Schutt, W. A. (2022). Demonic and Divine Attributions around COVID-19 Vaccines: Links with Vaccine Attitudes and Behaviors, QAnon and Conspiracy Beliefs, Anger, Spiritual Struggles, Religious and Political Variables, and Supernatural and Apocalyptic Beliefs. Religions, 13(6), 519. Retrieved from https://www.mdpi.com/2077-1444/13/6/519

Galbadage, T., Peterson, B. M., Wang, D. C., Wang, J. S., & Gunasekera, R. S. (2020). Biopsychosocial and Spiritual Implications of Patients With COVID-19 Dying in Isolation. Front Psychol, 11, 588623. doi:10.3389/fpsyg.2020.588623

Gonçalves Júnior, J., Moreira, M. M., & Rolim Neto, M. L. (2020). Silent Cries, Intensify the Pain of the Life That Is Ending: The COVID-19 Is Robbing Families of the Chance to Say a Final Goodbye. Front Psychiatry, 11, 570773. doi:10.3389/fpsyt.2020.570773

Hasin, D. S., O’Brien, C. P., Auriacombe, M., Borges, G., Bucholz, K., Budney, A., … Grant, B. F. (2013). DSM-5 criteria for substance use disorders: recommendations and rationale. Am J Psychiatry, 170(8), 834–851. doi:10.1176/appi.ajp.2013.12060782

Johnson, S. S. (2021). Addressing Mental Health and Substance Use Disorders Amid and Beyond the COVID-19 Pandemic. Am J Health Promot, 35(2), 299–301. doi:10.1177/0890117120983982a

Khadrawy, Y. A., Sawie, H. G., Abdel-Salam, O. M. E., & Hosny, E. N. (2017). Cannabis exacerbates depressive symptoms in rat model induced by reserpine. Behav Brain Res, 324, 41–50. doi:10.1016/j.bbr.2017.02.015

Koenig, H. G. (2009). Research on religion, spirituality, and mental health: a review. Can J Psychiatry, 54(5), 283–291. doi:10.1177/070674370905400502

Levine, R. L. (2022). Addressing the Long-term Effects of COVID-19. Jama, 328(9), 823–824. doi:10.1001/jama.2022.14089

Livne, O., Wengrower, T., Feingold, D., Shmulewitz, D., Hasin, D. S., & Lev-Ran, S. (2021). Religiosity and substance use in U.S. adults: Findings from a large-scale national survey. Drug Alcohol Depend, 225, 108796. doi:10.1016/j.drugalcdep.2021.108796

Lowe, G. B., Wang, D. C., & Chin, E. G. (2022). Experiential Avoidance Mediates the Relationship between Prayer Type and Mental Health before and through the COVID-19 Pandemic. Religions, 13(7), 652.

Marsiglia, F. F., Ayers, S. L., & Hoffman, S. (2012). Religiosity and adolescent substance use in central Mexico: exploring the influence of internal and external religiosity on cigarette and alcohol use. Am J Community Psychol, 49(1-2), 87–97. doi:10.1007/s10464-011-9439-9

Rhew, I. C., Cadigan, J. M., & Lee, C. M. (2021). Marijuana, but not alcohol, use frequency associated with greater loneliness, psychological distress, and less flourishing among young adults. Drug Alcohol Depend, 218, 108404. doi:10.1016/j.drugalcdep.2020.108404

Ross, S., & Peselow, E. (2012). Co-occurring psychotic and addictive disorders: neurobiology and diagnosis. Clin Neuropharmacol, 35(5), 235–243. doi:10.1097/WNF.0b013e318261e193

SAMHSA. (2021). 2020 National Survey on Drug Use and Health Public Use File Codebook. Center for Behavioral Health Statistics and Quality. Substance Abuse and Mental Health Services Administration (SAMHSA), Rockville, MD.

She, K. T. R., Wang, D. C., Canada, A. L., & Poston, J. M. (2022). The Impact of Financial Health on the Spiritual, Mental, and Relational Health of Christian Graduate Students. Pastoral Psychology. doi:10.1007/s11089-022-01031-1

Smith, C. L., Waters, S. F., Spellacy, D., Burduli, E., Brooks, O., Carty, C. L., … Barbosa-Leiker, C. (2022). Substance use and mental health in pregnant women during the COVID-19 pandemic. J Reprod Infant Psychol, 40(5), 465–478. doi:10.1080/02646838.2021.1916815

Smith, K., Wang, D., Canada, A., Poston, J. M., Bee, R., & Hurlbert, L. (2022). The biobehavioral family model with a seminarian population: A systems perspective of clinical care. Front Psychol, 13, 859798. doi:10.3389/fpsyg.2022.859798

Stewart, C. (2001). The influence of spirituality on substance use of college students. J Drug Educ, 31(4), 343–351. doi:10.2190/hepq-cr08-mgyf-yylw

Upenieks, L. (2022). Religious/spiritual struggles and well-being during the COVID-19 pandemic: Does “talking religion” help or hurt? Review of Religious Research, 64(2), 249–278. doi:10.1007/s13644-022-00487-0

Verdery, A. M., Smith-Greenaway, E., Margolis, R., & Daw, J. (2020). Tracking the reach of COVID-19 kin loss with a bereavement multiplier applied to the United States. Proc Natl Acad Sci U S A, 117(30), 17695–17701. doi:10.1073/pnas.2007476117

Weber, S. R., & Pargament, K. I. (2014). The role of religion and spirituality in mental health. Curr Opin Psychiatry, 27(5), 358–363. doi:10.1097/yco.0000000000000080

Weinberger, A. H., Pacek, L. R., Sheffer, C. E., Budney, A. J., Lee, J., & Goodwin, R. D. (2019). Serious psychological distress and daily cannabis use, 2008 to 2016: Potential implications for mental health? Drug Alcohol Depend, 197, 134–140.

Wolitzky-Taylor, K., Operskalski, J. T., Ries, R., Craske, M. G., & Roy-Byrne, P. (2011). Understanding and treating comorbid anxiety disorders in substance users: review and future directions. J Addict Med, 5(4), 233–247. doi:10.1097/ADM.0b013e31823276d7

Womack, S. R., Shaw, D. S., Weaver, C. M., & Forbes, E. E. (2016). Bidirectional Associations Between Cannabis Use and Depressive Symptoms From Adolescence Through Early Adulthood Among At-Risk Young Men. J Stud Alcohol Drugs, 77(2), 287–297. doi:10.15288/jsad.2016.77.287

